# Machine Learning Interpretability Methods to Characterize Brain Network Dynamics in Epilepsy

**DOI:** 10.1101/2023.06.25.23291874

**Authors:** Dipak P. Upadhyaya, Katrina Prantzalos, Suraj Thyagaraj, Nassim Shafiabadi, Guadalupe Fernandez-BacaVaca, Subhashini Sivagnanam, Amitava Majumdar, Satya S. Sahoo

## Abstract

The rapid adoption of machine learning (ML) algorithms in a wide range of biomedical applications has highlighted issues of trust and the lack of understanding regarding the results generated by ML algorithms. Recent studies have focused on developing interpretable ML models and establish guidelines for transparency and ethical use, ensuring the responsible integration of machine learning in healthcare. In this study, we demonstrate the effectiveness of ML interpretability methods to provide important insights into the dynamics of brain network interactions in epilepsy, a serious neurological disorder affecting more than 60 million persons worldwide. Using high-resolution intracranial electroencephalogram (EEG) recordings from a cohort of 16 patients, we developed high accuracy ML models to categorize these brain activity recordings into either seizure or non-seizure classes followed by a more complex task of delineating the different stages of seizure progression to different parts of the brain as a multi-class classification task. We applied three distinct types of interpretability methods to the high-accuracy ML models to gain an understanding of the relative contributions of different categories of brain interaction patterns, including multi-focii interactions, which play an important role in distinguishing between different states of the brain. The results of this study demonstrate for the first time that post-hoc interpretability methods enable us to understand why ML algorithms generate a given set of results and how variations in value of input values affect the accuracy of the ML algorithms. In particular, we show in this study that interpretability methods can be used to identify brain regions and interaction patterns that have a significant impact on seizure events. The results of this study highlight the importance of the integrated implementation of ML algorithms together with interpretability methods in aberrant brain network studies and the wider domain of biomedical research.

## 1. Introduction

Brain network dynamics play a critical role in various cognitive functions of the brain. Understanding changes in these network dynamics has been crucial in exploring disease mechanisms in complex neurological disorders such as epilepsy [1]. Epilepsy is a serious neurological disease, affecting over 60 million individuals worldwide. More than 40% of epilepsy patients suffer from seizures that cannot be controlled with traditional anti-epileptic medications. Epilepsy as a chronic disorder has significant detrimental effects on the quality of life (QoL) of patients and it imposes substantial economic burden together with an increased risk of mortality [2]. Therefore, the systematic characterization of changes in brain network dynamics before and during epileptic seizure has been a major focus of research studies, which aim to accurately predict seizures [3], precisely localize seizure onset zone to enhance surgical outcomes in patients refractory to medication[4], and identify biomarkers for poorly understood phenomena such as Sudden and Unexpected Death in Epilepsy (SUDEP) [5].

At present, we have limited understanding of the underlying principles that influence changes in interactions patterns of neurons that lead to the creation of coordinated complexes to generate seizure activities [2]. To address the limitations of existing computational methods that relied on graph-based models of binary brain interactions, recent studies have demonstrated the effectiveness of rigorous mathematical models based on algebraic topology [6]. Algebraic topology methods have successfully modeled higher-order brain interactions between multiple brain regions using a generalization of the graph structures called a *simplex*, which together with robust methods such as persistent homology enables the precise characterization of epilepsy networks. In our previous work [6], we have developed topological data analysis (TDA) methods to characterize seizure networks using a visual representation called a persistent diagram, which systematically tracks the formation and disintegration of high dimensional topological structures during different stages of a seizure. However, persistent diagrams are challenging to analyze using quantitative methods and they provide limited insights into the details of aberrant brain network dynamics in epilepsy patients.

## 2. Background

### 2.1 Framework for brain network analysis in epilepsy

To address this challenge, we developed a machine learning (ML) framework (**Figure 1**) for the analysis of topological structures derived from high-resolution EEG data recorded from refractory epilepsy patients who undergo pre-surgical evaluation at the University Hospitals Cleveland Medical Center epilepsy monitoring unit, which is a level 4 facility that regularly performs epilepsy surgery. The data is recorded using electrodes implanted in the brain using the stereotactic placement scheme to record EEG (SEEG) [7]. Intracranial EEG recordings have gained recognition as the “gold standard” for pre-surgical evaluation as the recordings provide high-quality signals recorded directly from brain regions, without interference from intermediate layers that often affect other types of recordings such as scalp EEG data [5]. These multi-channel signals correspond to electrical activity from different brain regions, and various methods have been developed to characterize the coupling between different signal channels during seizure-related events [8]. One widely used method for the computation of the nonlinear regression coefficient h^2^ by Pijn et al., which encapsulates the coupling between two EEG signals during seizure events. The nonlinear correlation measures are presented as a matrix (M_ij_) showing the signal channel coupling and directionality weights [9].

**Figure 1:**
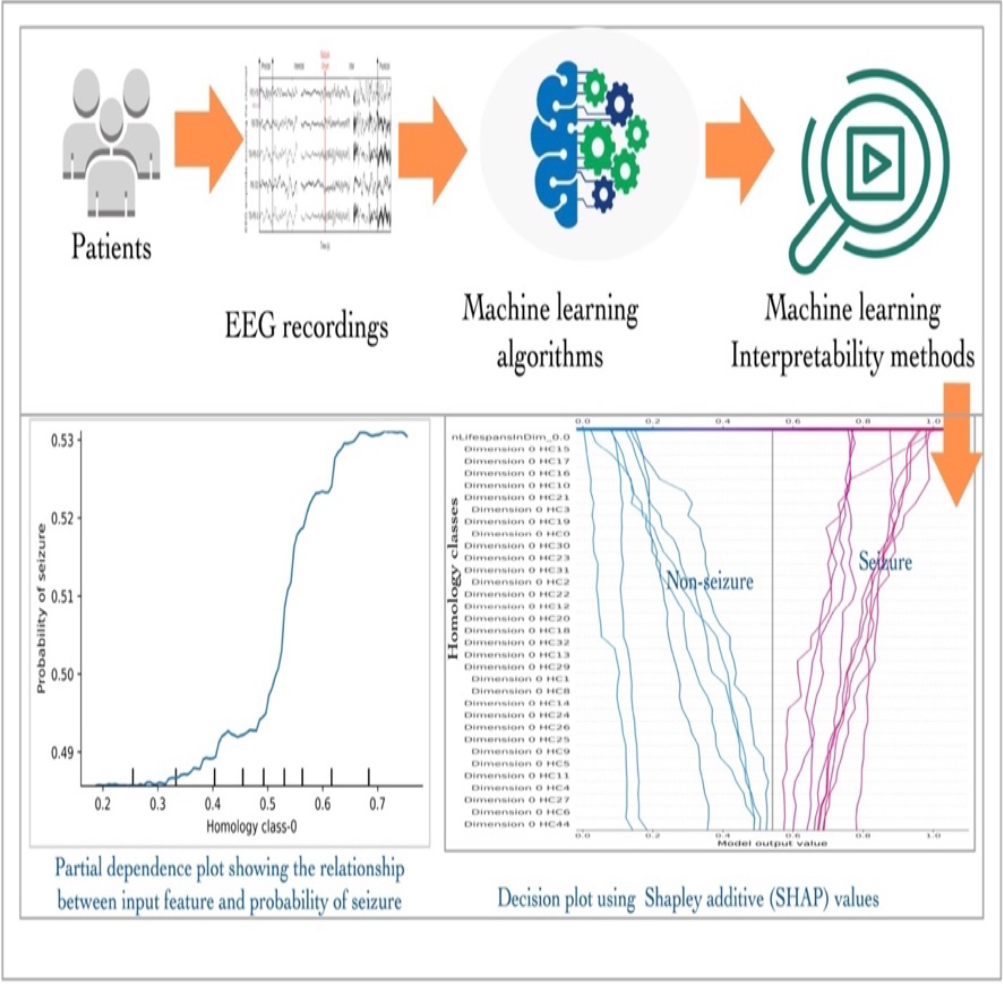
An overview of the machine learning interpretability framework developed within this study. In this work, we use Shapley Additive Value (SHAP) and Partial Dependence Plot (PDP) to emphasize the importance of brain interaction patterns in characterizing brain states.

In this study, we used topological structures called clique complexes, which were derived from coupling measures computed from SEEG recordings using a widely applied non-linear correlation coefficient (*h*^*2*^) [Pijn]. The clique complexes represent all-to-all connected components that are well-suited for studying co-activated brain structures during seizure events. Although ML algorithms have been used to analyze EEG data [10]; however, previous studies have relied only on signal processing methods to generate learning features for ML models, which limit their scalability and accuracy as it is difficult to represent interaction patterns across multiple electrode channels. Recent studies have employed ML algorithms to identify brain states and interaction patterns between different brain regions [11, 12], including pilot studies focused on using topological structures for classification of seizure and non-seizure brain states [13, 14]. Building on our previous studies and pilot studies performed by other groups, we have developed high accuracy classical and deep neural network (DNN) models for the following classification tasks:

□ *Task 1: Binary classification* of SEEG recordings during seizures (ictal events) and during baseline events with limited or no seizure related brain activities (e.g., sleep, awake)
□ *Task 2: Multiclass classification of* brain interaction patterns recorded during different stages of seizure propagation, including seizure onset, ictal phase 1 (first spread of seizure to additional brain regions), ictal phase 2, etc.

Following the development of ML algorithms that demonstrated high accuracy for the above two tasks, we implemented three interpretability methods, which were used to characterize the individual contributions of different categories of brain interaction patterns (represented by the corresponding clique complexes). The rest of the paper is organized as follows: In Section 4, we describe implementation of the ML algorithms for the binary and multiclass classification tasks and the interpretability methods for characterizing these classification analysis. In section 5, we discuss the results from this classification and subsequent interpretability analysis. Finally, in section 6 we provide discussion and conclusions from these studies.

## 3. Related Work

### Use of algebraic topology for characterizing epileptic seizure networks

Existing network analysis techniques, which utilize graph models where nodes symbolize brain structures and edges symbolize neural interactions, are constrained to modeling binary interactions between pairs of nodes. However, in neurological conditions such as epilepsy, it is common to witness simultaneous interactions involving three or more brain structures. As a result, Topological Data Analysis (TDA) methods, such as persistent homology capable of characterizing topological structures in noisy datasets, have garnered considerable interest in brain network studies. TDA allows data to be represented as a simplicial complex, an extension of a graph consisting of simplices that portray multidimensional interactions. A k-simplex refers to the convex hull encompassing k+1 interactive nodes, with the value of k representing the dimension of the simplex.

TDA analyzes simplicial complexes to identify topological structures, called homology classes, which are the boundary of a collection of k-simplices that form a closed structure (such as a loop or a tunnel) For a more in depth understanding of simplicial complex analysis, we direct the readers to the work of Edelsbrunner et. al [15]. The dimension of a homology class is equivalent to the dimension of the simplices forming the closed structure. A dimension 0 homology class represents the vertices in a network; a dimension 1 homology class represents the boundary of a 1-dimensional closed structure (e.g., line); a dimension 2 homology class represents the boundary of a 2-dimensional closed structure (e.g., a triangle), and higher dimensional homology classes represent multi-dimensional interaction structures[16]. Persistent homology is a TDA method that tracks the lifespan of these homology classes across different thresholds values (called filtration) and the lifespan of these homology classes, that is difference between the formation and termination of homology classes, can be used to describe the strength of interaction between brain regions. Homology classes with larger lifespans are assumed to describe more fundamental features of the complex; however, short-lived homology classes may be equally as important in characterizing networks [17]. In this work we use lifespans values of different homology classes as input features for ML algorithms and use interpretability methods to characterize the significance of specific homology classes across different dimensions in the two classification tasks.

## 4. Methods

### 4.1 Dataset and Patient Cohort

A cohort consisting of 16 patients was selected based on clinically determined inclusion exclusion criteria to identify patients who were considered for surgical intervention to disrupt epileptic seizure network. The study dataset included SEEG recordings for two seizure events in each patient. In addition to seizure event, recordings from non-seizure events (without seizure activity) were also selected for each patient and matched in duration to the longer of the two seizure events. Using the Neuro-Integrative Connectivity (NIC) tool [6], bidirectional network graphs for one-second epochs were computed using the h^2^ nonlinear correlation coefficient measure [9]. The NIC TDA module was used to compute persistent homology for each bidirectional network graph to determine the lifespan of homology classes in each one-second epoch. The lifespan values of all homology classes in each epoch were grouped together in a list and labeled with either seizure or non-seizure class label. These lists together with their class labels were used as input features for the various ML algorithms.

### 4.2 Data Augmentation for Imbalanced Data

High quality data, including a representative distribution of class labels, is an essential component of high-performance ML algorithm development process. In our study, the original dataset consisted of data with six distinct class labels with the class label, seizure *onset*, occurring in 294 epochs (9% of total class labels) class label *ictal 3*, occurred in 755 epochs (comprising 23% of the total class labels). Therefore, this skewed distribution of class labels resulted in an imbalanced dataset. To address this issue, we used the Synthetic Minority Oversampling Techniques (SMOTE) data augmentation method [18]. Imbalanced data, where the number of samples from each class differs significantly, is a common challenge in real-world scenarios such as healthcare. SMOTE uses randomized interpolation of neighboring minority labels from a set of instances representing the minority classes to generate new instances of minority classes, enhancing classifier generalizability. It does not introduce new information or variation but rather generates new data points using the K-nearest method from the feature space.

### 4.3 Machine Learning Models for binary and multi class classification

We used three ML algorithms in this study, that is Support Vector Machine (SVM), Random Forest (RF), and Deep Neural Network (DNN), to distinguish between different brain states and understand the interactions between different brain regions. To implement the SVM model, we used the scikit-learn package with hyperparameters values selected using the automated grid search method (C = 10, kernel type = poly). The DNNmodel was, implemented using TensorFlow Keras API with five hidden layers with a varying number of nodes using the rectified linear unit (ReLU) activation. We employed the “rmsprop” optimizer, “Binary_crossentropy” loss, and a learning rate of 0.001. The final output layer used sigmoid activation for binary classification and softmax for multiclass. Random Forest, used for ensemble learning, employed RandomForestClassifier from scikit-learn with specific parameters (n_estimators = 100, max_depth = 5, min_samples_leaf = 1, min_samples_split = 15). Hyperparameter optimization utilized the Keras grid search library. The primary objective of the three ML models consisted of achieving high accuracy results for binary as well as multiclass classification tasks and subsequently the use of ML interpretability methods to characterize the contribution of homology classes across dimensions to the accuracy results.

### 4.4 Interpretability methods

In the second phase of the study, we implemented three model agnostic ML interpretability methods: Shapley Additive Value (SHAP), Permutation Feature Importance (PFI) and Partial Dependence Plots (PDP) [19-21]. In this study, we distinguish between ML interpretability methods, which characterizes the changes in results of ML algorithm based on variation of input features, which characterize the dynamics of the ML model [22]. The SHAP method, grounded in cooperative game theory, offers comprehensive insights into the importance of individual features for a model, and illustrates how each feature contributes to individual predictions. The SHAP features importance and SHAP decision plot were employed as interpretability method, enabling visualization of the result derived from machine learning models. These interpretability methods provide insights into the model’s decision-making process and the impact of different input features on the predictions. The PFI method was also used, as it is more computationally efficient than SHAP. PFI allows the evaluation of feature importance by manipulating the value of individual features and observing the subsequent change in model performance. This method gave us with further insights regarding the of relative importance of each feature in the predictive process.

In addition to the two ML interpretability methods, we also charted partial dependency plots (PDP) to visually characterize the performance of the ML algorithms as the value of other features of interest were varied. PDP enable users to characterize feature importance, linearity, interactions, and potential biases of input features and enhancing transparency as well as understanding of model behavior. These three interpretability methods provide insights into the model’s decision-making process and impact of different input features on the predictions.

## 5. Results

We utilized a standardized approach to evaluate the classification performance. We used the Random Forest ML model with the highest accuracy values of 0.88 and 0.70 for binary and multiclass classification, respectively for implementation of the ML interpretability methods. We applied interpretability methods to identify important features in classifying seizure and non-seizure using homology classes in multiple dimensions. By identifying important homology classes per dimension, the interpretability method provides insights into which features of the data are most relevant for distinguishing between seizure and non-seizure states. For multiclass classification, interpretability methods help understand the mechanism of how the model is identifying different stages of seizure propagation. This helps to understand which dimension of homology classes the model uses to differentiate between different stages of seizures.

**Table 1:** Mean accuracy values of binary and multiclass classification across all three algorithms for the complete study cohort (95% Confidence interval)

| **Machine learning algorithms** | **Binary Classification**<br>(Seizure and non-seizure states) | **Multiclass classification**<br>(States of seizure progressions) |
| --- | --- | --- |
| Support Vector Machine | 0.78(0.76-0.80) | 0.62(0.58-0.64) |
| Deep Neural Network | 0.80(0.78-0.82) | 0.60(0.57-0.63) |
| Random Forest | 0.88 (0.86-0.89) | 0.70(0.67-0.73) |

### 5.1. Interpretability results for binary classification task

We first applied interpretability methods to ascertain the contribution of homology classes and their lifespans to the classification of seizure and non-seizure states in a Random Forest model. We show that the lifespan of homology classes in dimension 0, representing the filtration (strength of correlation) at which an electrode becomes involved in any sort of connection (pairwise or multidimensional), are consistently the most distinguishing features in seizure or non-seizure classification in single patient analyses or across all patients (**Figure 2**). We also show that there is some importance of the dimension 1 homology class lifespans in both SHAP and PFI methods.

**Figure 2:**
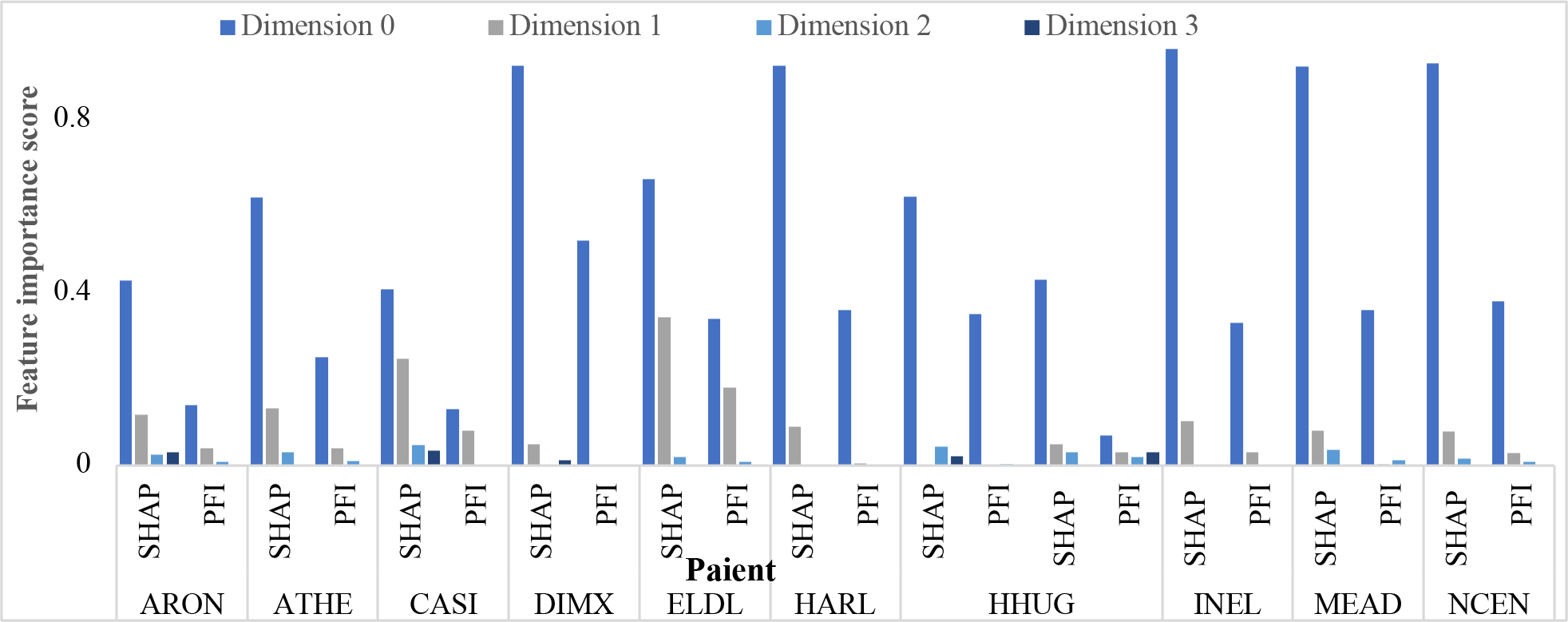
Feature importance scores for homology classes along each dimension calculated using SHAP and PFI (to space constraints, we present only 10 patients results)

A homology class in dimension 1 is the boundary of a collection of 1-simplices (pairwise connections) that form a closed path (such as a loop or a cycle). for each dimension 1 homology class, there is a set of electrodes for which there is a continuous and closed path of pairwise interaction; however, at least one pair of electrodes in the interacting set does not directly interact. The lifespan of a dimension 1 homology class thus represents how much more interaction is needed for an existing path of pairwise interactions to interact with other electrodes. Our results indicate that the lifespans of these lower-dimensional interactions are the most influential in classifying seizure and non-seizure states in both feature importance methods, SHAP and PFI.

To identify the impact of different homology classes on the classification of seizure and non-seizure events in EEG samples, we selected the first one-second epoch from seizure and non-seizure periods from a randomly selected patient and visualized them using a SHAP decision plot (**Figure 3**). In this plot, homology classes are labeled with their dimension and their order in the lifespan list (homology class 1 (HC1), HC2, etc.), and are organized in decending order of their importance as determined by their SHAP values. The features depicted in Figure 3 are arranged in a descending sequence based on the feature importance scores calculated by the SHAP methodology. SHAP values show the difference between the expected and the actual outputs of a machine learning algorithm. Consequently, the decision plots for both patients begin at the base of the y-axis, corresponding to the shown result determined by the machine learning algorithm. In the subsequent process, the SHAP value associated with each feature is cumulatively added to this expected value, culminating in the final classification output produced by the machine learning algorithm.

**Figure 3:**
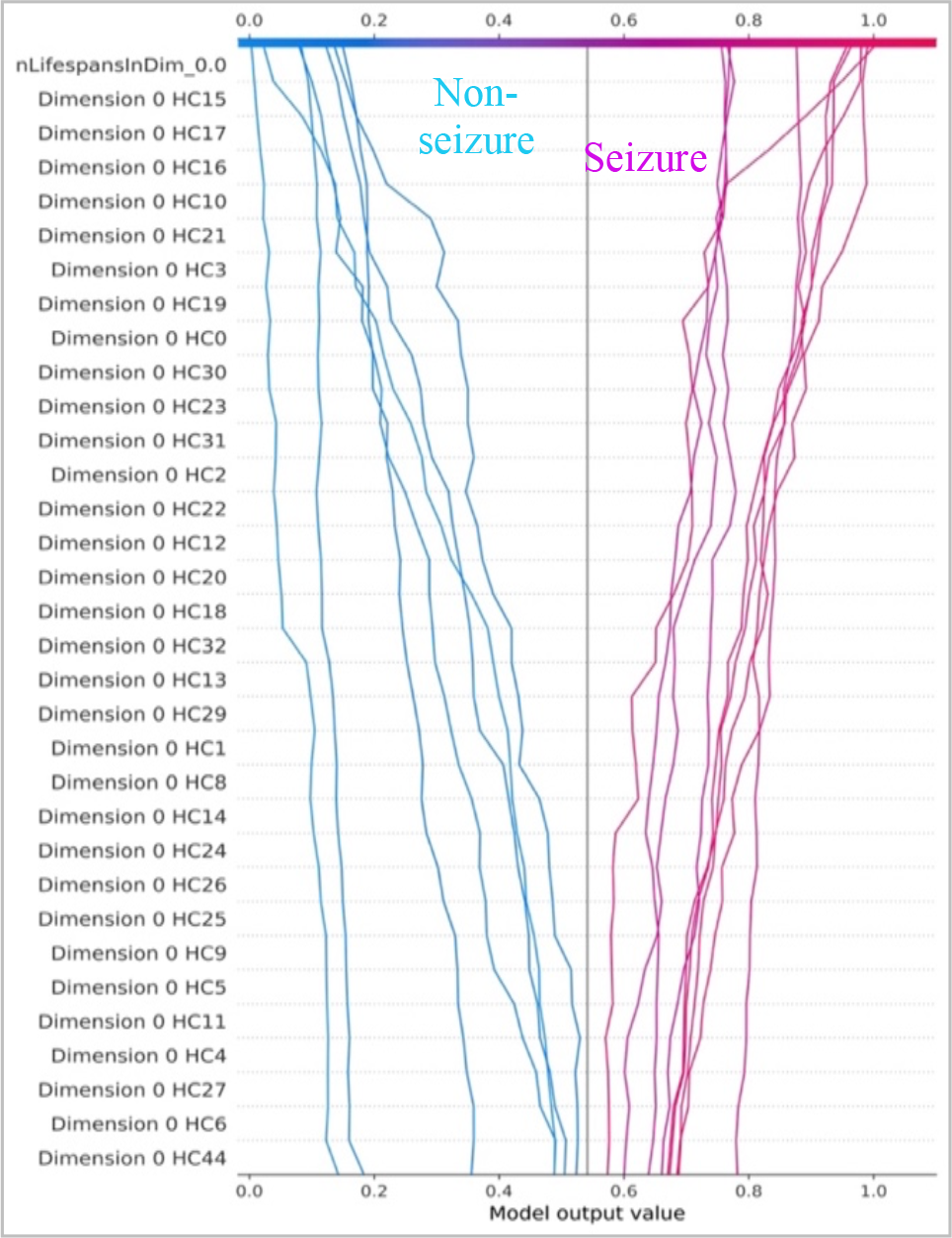
A decision plot visualization of SHAP feature importance score computed for different homology classes across different dimensions.

The findings illustrated in **Figure 3** highlight that a significant number of homology classes in dimension 0 play a crucial role in accurately classifying seizure and non-seizure events. This further emphasizes the importance of lower dimensional homology classes and their lifespans in characterizing seizure and non-seizure states. Through **Figure 3**, we are able to identify several dimension 0 homology classes which are most influential in classifying seizure and non-seizure states, such as homology class 15, 16, and 17 (identifiers generated by the GUDHI tool). The importance of the difference in lifespan values for these homology classes can be seen further when looking at persistence diagrams of each of the corresponding one-second epochs (**Figure 4**). For example, using the same epochs as selected for SHAP decision plot visualization, we suggest that asleep non-seizure periods might be classified by recognizing higher lifespans for homology classes 15, 16, and 17 compared to seizure onset; however, awake non-seizure periods might be characterized by much lower lifespans for homology classes 15, 16, and 17 (identifiers generated by the GUDHI tool).

**Figure 4:**
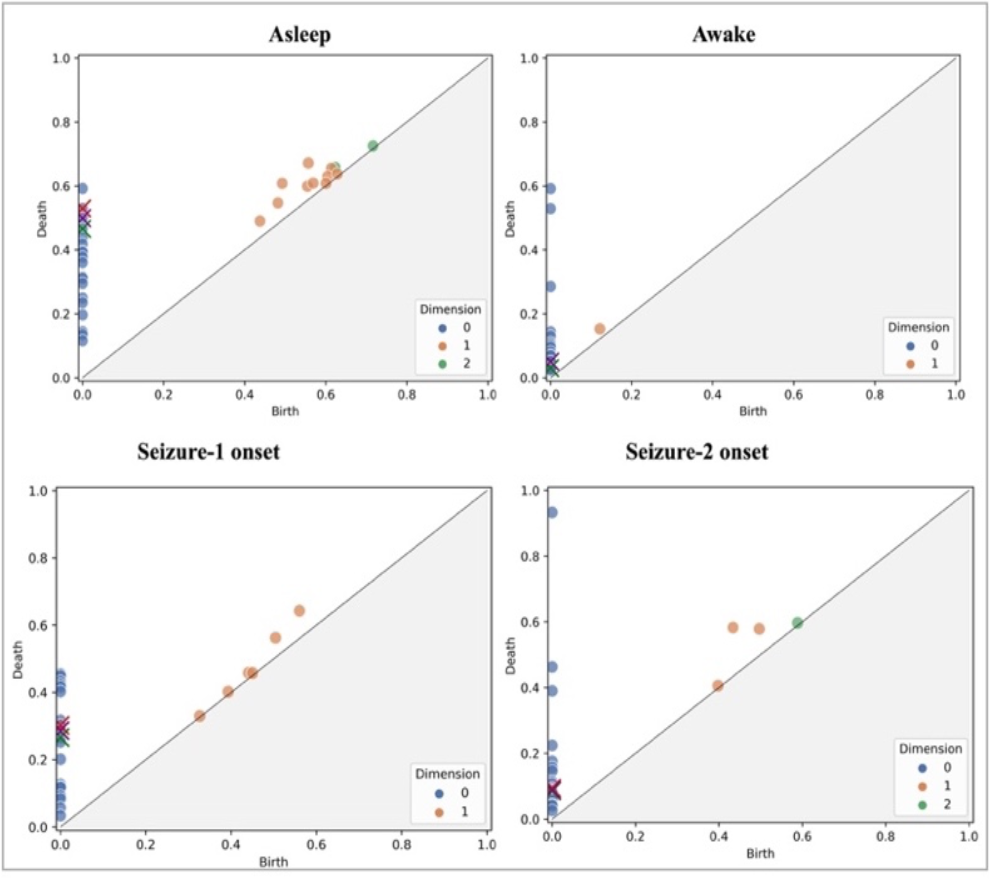
Scatter plots of three most distinguishing homology classes for a patient (persistence diagrams) corresponding to asleep, awake, seizure one onset, and seizure two onset states. Homology classes marked by colors red, green, and purple in dimension 0 were the most influential features.

In addition to feature importance score, we also applied PDP to our random forest model. PDP illustrates the functional relationship between homology classes, dimensions, and seizure probability. While SHAP identified dimension 0 homology classes to be the most essential for classification, PDP reveals some relationships in higher dimensions to have significant impact on seizure classification (**Figure 5**). Our results show that dimension 0 homology class 0, dimension 1 homology class 1, and dimension 5 homology class 0 have a linear effect on seizure probability, with increasing dimensions and classes lifespan values corresponding to higher probabilities. However, dimension 2 homology class 2, dimension 3 homology class 1, and the maximum dimension exhibit nonlinear effects. The relationship between these features and seizure probability does not follow a linear trend, but rather shows fluctuations at specific threshold values. In **Figure 5**, we observe an initial increase in seizure probability as the lifespan value of dimension 2 homology class 0 reaches 0.03, followed by a substantial decrease beyond this threshold. These findings emphasize the importance of considering both dimension and homology class in analyzing EEG recordings and highlight the complex nature of their relationship.

**Figure 5:**
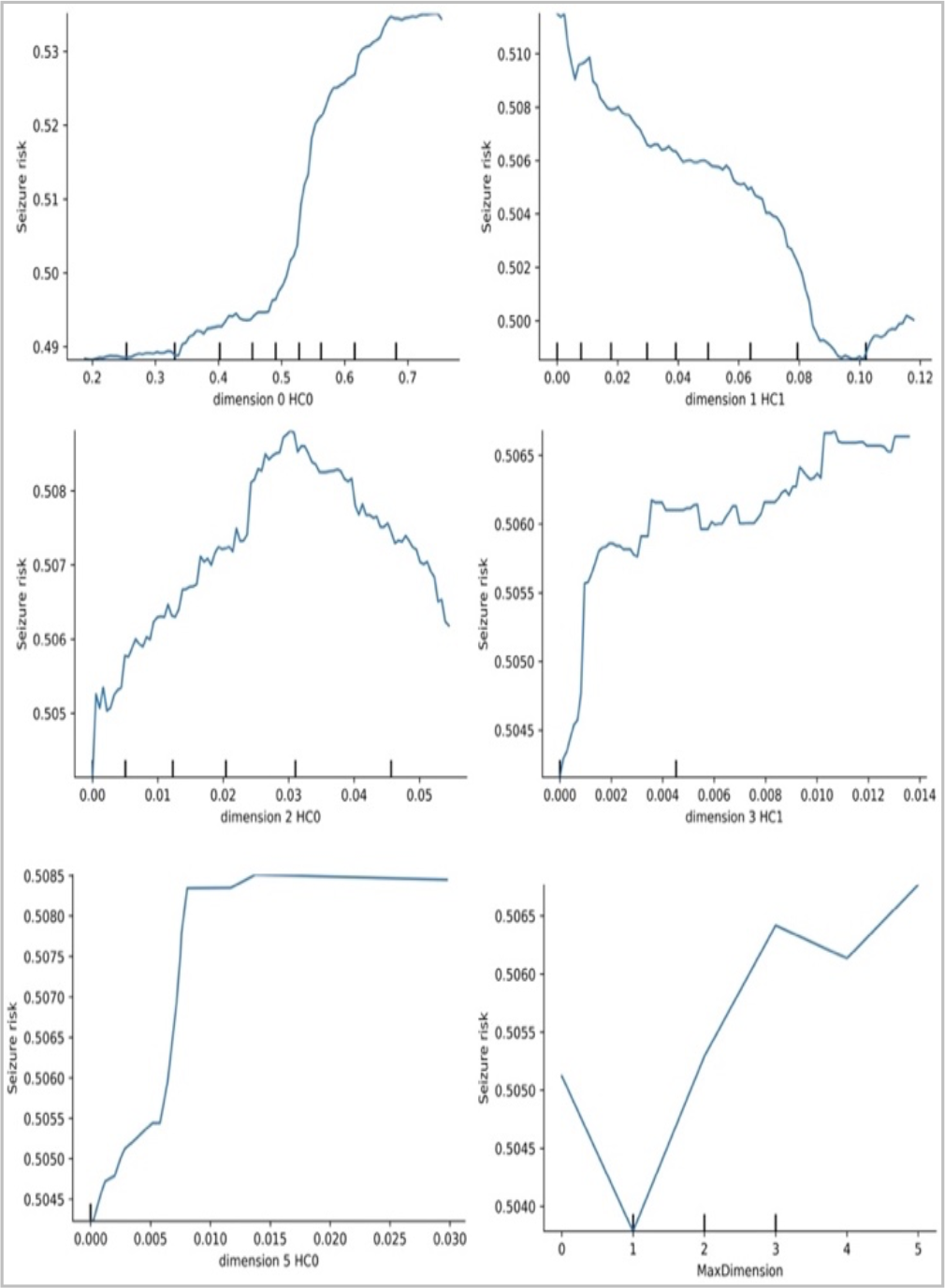
The partial dependence plot (PDP) for five homology classes with highest feature importance scores together with partial dependency score of the number of dimensions.

### 5.2. Interpretability results for multiclass classification task

In order to interpret the random forest algorithm results for our multiclass classification of seizure periods, we computed feature importance score using SHAP and PFI (**Figure 6**) to demonstrate the feature importance scores of homology classes in each dimension. Similar to binary classification results, our multiclass classification results reveal that lifespans of homology classes in dimension 0 (representing the lowest filtration at which an electrode interacts with any other electrodes) make significant contributions to classification of the seizure periods in both SHAP and PFI interpretability methods.

**Figure 6:**
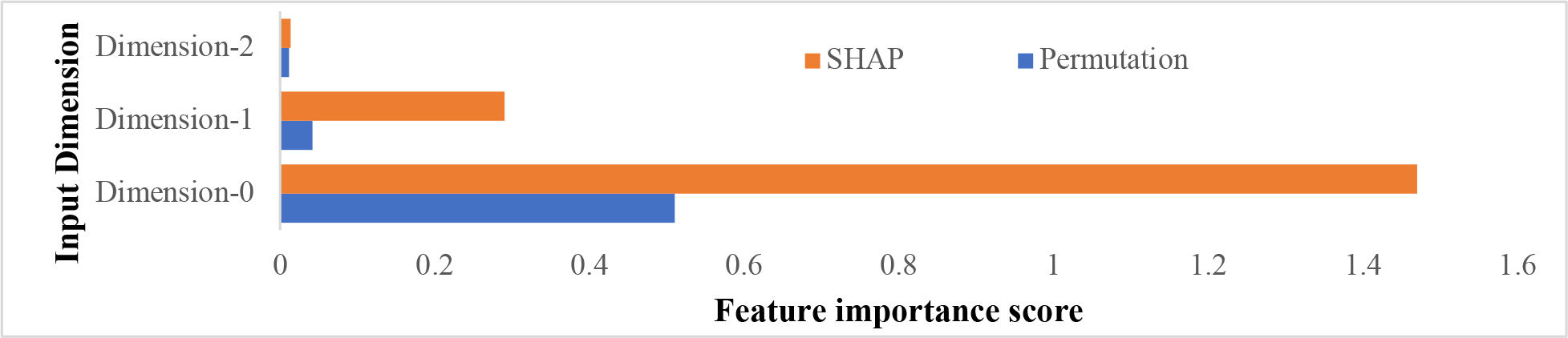
Feature importance scores calculated using SHAP and permutation methods for multiclass classification with RF algorithm.

## 5. Discussion and Conclusions

The results of the study demonstrate that ML interpretability methods can effectively identify brain network connections involved in seizure events. This study provides new avenues for understanding complex brain network dynamics in epilepsy and other disorders, highlighting the potential of ML interpretability methods to provide insights into neurological disorders and brain network dynamics. The lifespan of homology classes in dimension 0 emerged as a consistently distinguishing feature in seizure and non-seizure classification. The importance of dimension 0 homology classes is further confirmed through SHAP decision plots, which underscore their role in accurately classifying seizure and non-seizure events. The life span of lower dimensional homology classes was, hence, the most influential in this classification. For the multiclass classification task, the feature importance score analysis again emphasizes the vital contribution of lifespans of dimension 0 homology classes. This study demonstrates the utility of ML interpretability methods such as SHAP and PFI to identify the significant features has been underscored. Our findings emphasize the importance of continued research into the integration of ML algorithms and interpretability methods in medical disciplines, enabling more accurate and personalized diagnosis and treatment options for patients. This study also contributes to the growing body of knowledge regarding the use of ML in medical applications, providing new avenues for understanding complex brain network dynamics in epilepsy and other disorders.

## Data Availability

The individual patient records cannot be made publicly available due to regulatory reasons.

